# COVID-19 and HIV co-infection: a living systematic evidence map of current research

**DOI:** 10.1101/2020.06.04.20122606

**Authors:** Gwinyai Masukume, Witness Mapanga, Doreen S van Zyl

## Abstract

The world currently faces two ongoing devastating pandemics. These are the new severe acute respiratory syndrome coronavirus 2/coronavirus disease 2019 (SARS-CoV-2/COVID-19) and the prior human immunodeficiency virus/acquired immune deficiency syndrome (HIV/AIDS) pandemics. The literature regarding the confluence of these global plagues expands at pace. A systematic search of the literature considering COVID-19 and HIV co-infection was performed.

After five months, from the beginning of the COVID-19 pandemic, there were at least thirty-five studies reported from thirteen countries. These ranged from individual case reports and series to cohort studies. Based on studies that could be extrapolated to the general population, co-infected individuals with suppressed HIV viral loads did not have disproportionate COVID-19 sickness and death. At least four patients, newly diagnosed with HIV recovered from COVID-19. Current evidence suggests that co-infected patients should be treated like the general population.

This ongoing living systematic evidence map of contemporary primary SARS-CoV-2 and HIV co-infection research provides a platform for researchers, policy makers, clinicians and others to more quickly discover and build relevant insights.

## Introduction

The confluence of the ongoing new severe acute respiratory syndrome coronavirus 2/coronavirus disease 2019 (SARS-CoV-2/COVID-19) and prior human immunodeficiency virus/acquired immune deficiency syndrome (HIV/AIDS) pandemics presents profound challenges for public health.[1, 2] On 31 December 2019, a pneumonia cluster of unknown cause was reported to the World Health Organization.[3] This report foreshadowed the emergence of a new pandemic. Due to the emergence of the COVID-19 pandemic, scientific research has been growing very fast.[4] We thus sought to provide an updated systematic curation and key insights from literature on co-infection with the pandemic viruses.

## Methods

Two independent reviewers (GM and WM) searched the following databases: PubMed (via the PubMed/MEDLINE interface), the Scientific Electronic Library Online, Scopus (via the EBSCO interface), African Journals OnLine, and the pre-print server MedRxiv (using key words).

The Boolean search approach, for 2020 literature, (COVID* OR SARS-CoV-2 OR severe acute respiratory syndrome OR coronavirus OR corona virus) AND (HIV OR AIDS OR human immunodeficiency virus), was used for each database (PubMed search strategy). The last online database searches were on 3rd June 2020. Searches were supplemented by a hand search of references and by use of the ‘cited by’ feature of Google Scholar. Inclusion criteria included primary studies reporting individuals with co-infection. Exclusion criteria included editorials, commentaries and reviews.

All records that were identified from the searches were collated into EndNote software. Duplicates were removed. The screening of titles, abstracts and full-texts were performed, guided by the criteria described earlier. An HIV clinician (DSvZ) acted as the third reviewer and provided domain specific insights.

## Results

Presented in order of the first published paper from a country, there were thirty five papers from thirteen countries: China,[5–11] Spain,[12–14] Italy,[15, 16] United States,[17–29] United Kingdom,[30,31] Turkey,[32] Germany,[33] South Africa,[34] Singapore,[35] Uganda,[36], Cyprus,[37] (Figure 1), Japan,[38] and Austria.[39] The first known case of COVID-19 and HIV co-infection was reported from China on 11 March 2020.[5] From then onwards, the number of reports has increased quickly (Figure 2). Please see the supplementary appendix for further details and summaries of the papers.

**Figure 1.**
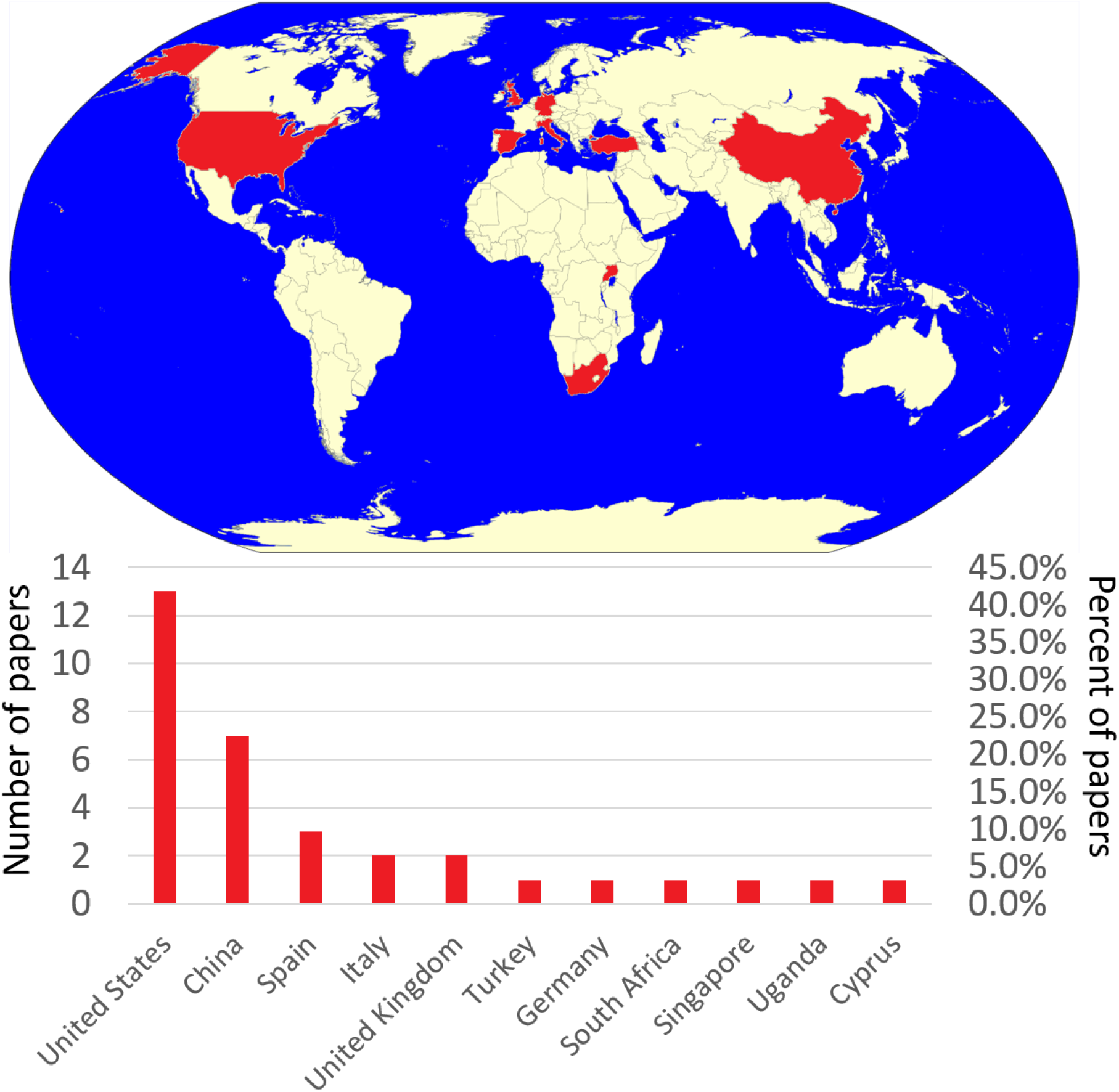
Countries reporting severe acute respiratory syndrome coronavirus 2 (SARS-CoV-2) and human immunodeficiency virus (HIV) co-infection up to 31 May 2020.

**Figure 2.**
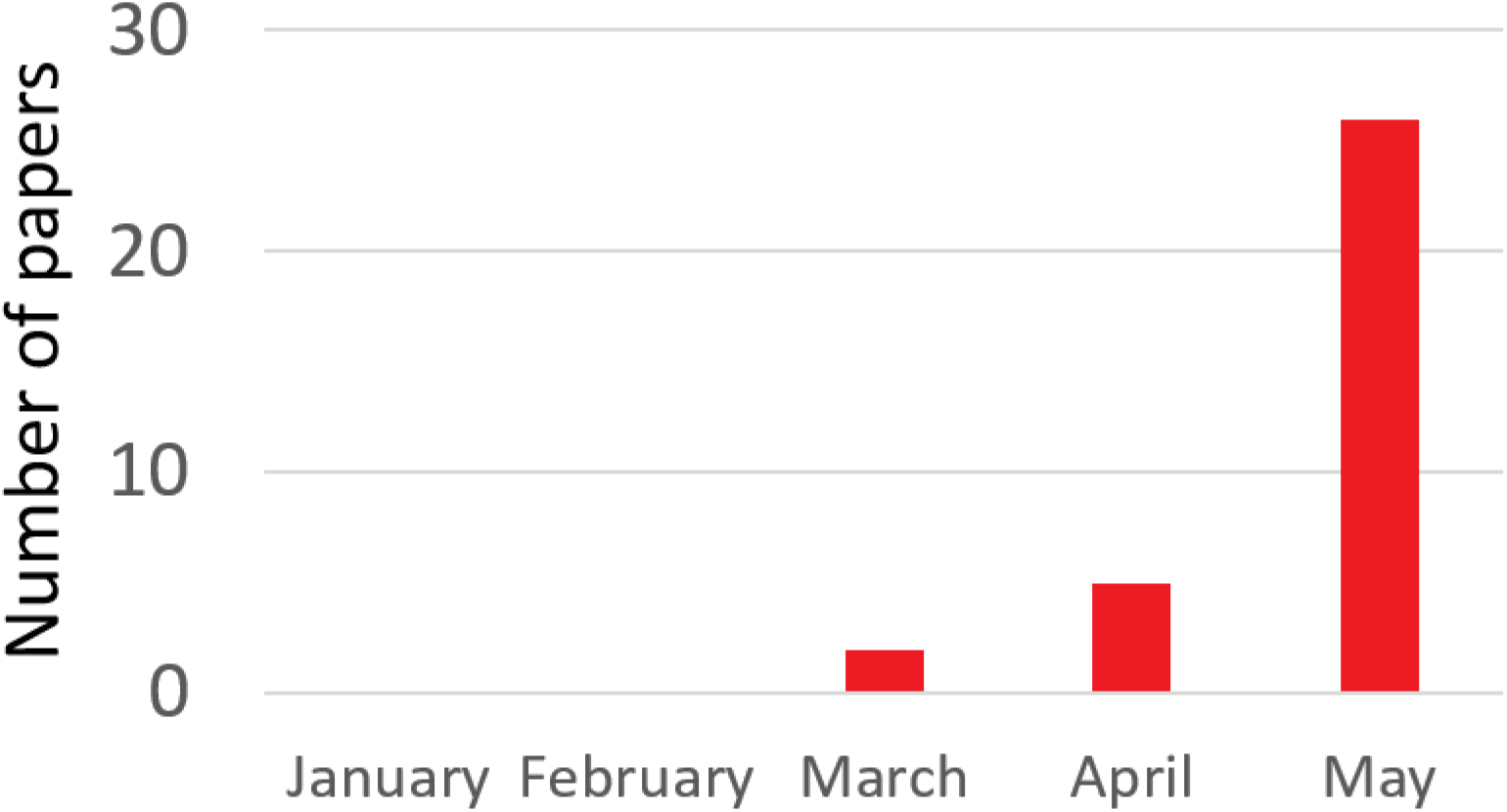
Monthly number of papers reporting severe acute respiratory syndrome coronavirus 2 (SARS-CoV-2) and human immunodeficiency virus (HIV) co-infection up to 31 May 2020.

Common symptoms included cough, fever, shortness of breath and fatigue. Loss of the sense of smell and taste as well as gastrointestinal symptoms like diarrhoea were not uncommon; however, there was a report of a patient with seizure activity.[20] Reports that were more generalisable to the broader population found no increased risk of HIV virally suppressed individuals being hospitalised or suffering excess COVID-19 morbidity and mortality.[28–30, 33, 40] Where there was increased risk of severe disease, patients had co-morbidities like hypertension, diabetes mellitus, obesity and chronic kidney disease.[31] Also, it was suggested that HIV-infected individuals ought to receive generally the same treatment for COVID-19 as the general populace.[14] At least four COVID-19 patients newly diagnosed with HIV recovered.[9, 10, 12, 38] The protease inhibitor, darunavir, was found unlikely to prevent SARS-CoV-2 infection in HIV patients.[15] There have been reports of co-infected patients with solid organ transplants of the kidney and liver recovering.[23, 39]

## Discussion

This brief evidence map provides an up-to-date overview of the SARS-CoV-2/COVID-19 and HIV/AIDS co-infection literature. Its strengths include the use of explicit search criteria and the provision of a supplementary appendix providing key summaries of the studies on the topic. Although there was no search language restriction, only English language studies were retrieved, which raises the possibility of bias.

The finding that darunavir is likely to be ineffective at preventing COVID-19 in people living with HIV is in keeping with the finding that lopinavir-ritonavir did not provide benefit in the treatment of COVID-19.[41] This suggests that existing HIV therapy regimens should not be switched in patients with SARS-CoV-2 co-infection to protease inhibitor based medication.

It is interesting to note that reports emerged from countries in a similar temporal sequence in concert with how the COVID-19 pandemic epicentre moved from Asia, then to Europe and then to the Americas. The current global burden of COVID-19 approximates, to an extent, the amount of co-infection literature from the respective continents. To the best of our knowledge, there are no reported cases of co-infection from Oceania, which has the lowest number of COVID-19 cases. Africa which has a substantial burden of HIV/AIDS, but a relatively low COVID-19 burden has reports of two patients.[1, 34, 36]

The interaction with opportunistic infections like tuberculosis and pneumocystis jirovecii merits further consideration.[12] As therapies for COVID-19 emerge, drug interactions mandate exploration. Along with mental health aspects associated with co-infection. Pregnant women represent a special population.[42] As far as we know, no co-infection case has been reported during pregnancy. Because literature is expanding at pace, this synthesis is envisaged to be living and will be updated regularly and with more detail. Due to the rapidly expanding literature [4] such brief evidence synthesis pieces provide a platform for researchers, policy makers, clinicians and others to quickly discover and build relevant insights.

## Conclusion

Based on studies that could be extrapolated to the general population, co-infected individuals with suppressed HIV viral loads did not have disproportionate COVID-19 morbidity and mortality. At least four patients, newly diagnosed with HIV survived COVID-19.

## Data Availability

Publicly available data

## Conflict of interest

None declared

## Ethics

Not required because publicly available data was used

## Funding

None

## Notes

### Competing Interest Statement

The authors have declared no competing interest.

### Author Declarations

Not required because publicly available data was used

